# Metabolic alkalosis and mortality in COVID-19

**DOI:** 10.1101/2022.04.01.22273291

**Authors:** Zhifeng Jiang

**Affiliations:** Xiaogan Hospital Affiliated to Wuhan University of Science and Technology; No.6, Square street, Xiaonan District, Xiaogan City, Hubei Province, China

**Keywords:** COVID-19, Metabolic alkalosis, mortality

## Abstract

**Background:** As a new infectious disease affecting the world, COVID-19 has caused a huge impact on countries around the world. At present, its specific pathophysiological mechanism has not been fully clarified. We found in the analysis of the arterial blood gas data of critically ill patients that the incidence of metabolic alkalosis in such patients is high.

**Method:** We retrospectively analyzed the arterial blood gas analysis results of a total of 16 critically ill patients in the intensive ICU area of Xiaogan Central Hospital and 42 severe patients in the intensive isolation ward, and analyzed metabolic acidosis and respiratory acidosis. Metabolic alkalosis and respiratory alkalosis, and the relationship between metabolic alkalosis and death.

**Result:** Among the 16 critically ill patients, the incidence of metabolic alkalosis was 100%, while the incidence of metabolic alkalosis in severe patients was 50%; the mortality rate in critically ill patients was 81.3%, and 21.4% in severe patients; The mortality of all patients with metabolic alkalosis is 95.5%,and 4.5% in without metabolic alkalosis.

**Conclusion:** The incidence of metabolic alkalosis in critically ill COVID-19 patients is high, and it is associated with high mortality.

## Introduction

COVID-19 has now swept the world, causing huge challenges and disasters to the global health system. At present, its detailed pathophysiological mechanism has not yet been fully clarified. The current research involves direct virus attack, humoral and cellular immunity, and nervous system damage. Endocrine disorders, respiratory and circulatory disorders, coagulation dysfunction and other aspects(1). There is still a lack of effective treatments. Critically ill patients still have a high mortality rate. Early data from Wuhan showed that the mortality rate of severely ill patients with COVID-19 was 62%, and the mortality rate of patients requiring mechanical ventilation was 81%(2). This manuscript analyzes the blood gas analysis data and deaths of critically ill patients in Xiaogan Central Hospital in March 2020, and finds that the incidence of metabolic alkalosis in critically ill patients is very high, and it is accompanied by a higher mortality rate.

## Method

Follow the Helsinki Declaration as revised in 2013,we analyzed 44 patients in the intensive isolation ward of Xiaogan Central Hospital, with an average age of 53 years, 27 males and 15 females. There was no history of Gitelman and Bartter syndrome in all patients, and exclude 2 cases of primary aldosteronism in patients. According to the diagnostic criteria of the fifth edition of China’s new coronavirus diagnosis and treatment guidelines (meet any of the following 1. Respiratory distress, RR>30 beats/min; 2. In the resting state, the oxygen saturation is <93%; 3. Arterial partial pressure of oxygen (PaO2)/inhaled oxygen concentration (FiO2) <300mmHg). All 42 patients were diagnosed as severe. Analyzing the arterial blood gas analysis data and death data of 42 patients; According to the same diagnostic criteria, we analyzed the arterial blood gas analysis data and death data of a total of 16 critically ill patients,11 were males and 5 were females, with an average age of 67 years. (meet one of the following conditions:1. Respiratory failure occurs and mechanical ventilation is required; 2.Shock;3. combined with other organ failure, ICU monitoring and treatment is required) in the intensive ICU of our hospital.

Analyze the arterial blood gas data of all patients, select the highest bicarbonate value as the statistical data, including carbon dioxide partial pressure (PaCO2), oxygen partial pressure (PaO2), bicarbonate (HCO3-), alkali excess (BE), serum potassium and calculate acid-base imbalance types, including metabolic acidosis, respiratory acidosis, metabolic alkalosis, respiratory alkalosis, respiratory acidosis combined with metabolic alkalosis, and analyze the mortality of critical and severe patients, at the same time, compare the mortality of patients with metabolic alkalosis and non-metabolic alkalosis. In addition, respiratory acidosis combined with metabolic alkalosis and metabolic alkalosis were combined as metabolic alkalosis, and the incidence of alkalosis and mortality were compared again, simultaneously compare the serum potassium of the two groups of patients. Use spss25.0 statistical software to analyze this data. The basic description of the count data is expressed by frequency and composition ratio, and the analysis of the difference between the two groups of count data uses the χ^2^ test, t test is used for measurement data, P<0.05 indicates that the difference is statistically significant.

## Result

There were 10 cases of acid-respiratory and metabolic alkalosis in critically ill patients, with an incidence rate of 62.5%, and 11 cases of acid-respiratory and metabolic alkalosis in severe patients, with an incidence of 26.2%, χ^2^ was 6.613, P=0.010, there was a statistical difference in the incidence of the two groups. The incidence of acid and alkali substitution in critical cases was significantly higher than that in severe cases. There were 6 cases of metabolic alkalosis alone in critically ill patients with an incidence rate of 37.5%, and 10 cases of metabolic alkalosis in severe patients with an incidence rate of 23.8%, χ^2^ was 1.087, P=0.297, there was no statistical difference in the occurrence of metabolic alkalosis between the two groups. However, when the number of cases of respiratory acidosis combined with metabolic alkalosis and metabolic alkalosis are combined, the incidence of metabolic alkalosis in critical cases is 100%, and the incidence of metabolic alkalosis in severe patients is 50%.(Table 1)

**Table1.** Comparison of the incidence of acid-base balance disorders in critically ill and critically ill patients.(AC &MA: Respiratory acidosis combined with metabolic alkalosis)

| group | case | critically ill | Severe | $\chi^2/t$ | P |
| --- | --- | --- | --- | --- | --- |
| AC &MA |  |  |  | 6.613 | 0.010 |
| Yes | 21 | 10(62.5%) | 11(26.2%) |  |  |
| No | 37 | 6(37.5%) | 31(73.8%) |  |  |
| Metabolic alkalosis |  |  |  | 1.087 | 0.297 |
| Yes | 16 | 6(37.5%) | 10(23.8%) |  |  |
| No | 42 | 10(62.5%) | 32(76.2%) |  |  |
| Death |  |  |  | 17.611 | 0.000 |
| Yes | 22 | 13(81.3%) | 9(21.4%) |  |  |
| No | 36 | 3(18.8%) | 33(78.6%) |  |  |
| Combined metabolic alkalosis |  |  |  | 12.541 | 0.000 |
| Yes | 21 | 16(100.0%) | 21(50.0%) |  |  |
| No | 21 | 0(0.0%) | 21(50.0%) |  |  |
| Serum potassium | - | $3.41 \pm 0.40$ | $3.68 \pm 0.46$ | -2.026 | 0.048 |

Comparing the two groups of patients with simple metabolic alkalosis and respiratory acidosis combined with metabolic alkalosis, it was found that among the dead patients, 14 cases of respiratory acidosis combined with metabolic alkalosis accounted for 63.6%, there are no respiratory acidosis combined with metabolic alkalosis in 8 case, accounting for 3.4%, with a χ^2^ of 11.546 and a P value of 0.001; When analyzing the death of patients with simple metabolic alkalosis, it was found that the death had nothing to do with simple metabolic alkalosis, χ^2^ was 0.318, P=0.573,when respiratory acidosis combined with metabolic alkalosis and metabolic alkalosis are combined as the number of cases of metabolic alkalosis, a total of 21 deaths, a ratio of 95.5%, and no metabolic alkalosis is 1 death, accounting for 4.5%, the χ^2^ was 15.383, and the P value is 0.000, the incidence of metabolic alkalosis is higher in deceased patients; Serum potassium in the critically ill was 3.41+-0.4mmol/L, and 3.68±0.46mmol/l in severe group, critically ill patients have lower blood potassium than severe patients (Table2).

**Table 2.** Number of deaths in acid-base imbalance.(AC &MA: Respiratory acidosis combined with metabolic alkalosis)

| group | case | no death | death | $\chi^2$ | P |
| --- | --- | --- | --- | --- | --- |
| AC &MA |  |  |  | 11.546 | 0.001 |
| Yes | 21 | 7(19.4%) | 14(63.6%) |  |  |
| No | 37 | 29(80.6%) | 8(36.4%) |  |  |
| metabolic alkalosis |  |  |  | 0.318 | 0.573 |
| Yes | 16 | 9(25.0%) | 7(31.8%) |  |  |
| No | 42 | 27(75.0%) | 15(68.2%) |  |  |
| Combined metabolic alkalosis |  |  |  | 15.383 | 0.000 |
| Yes | 37 | 16(44.4%) | 21(95.5%) |  |  |
| No | 21 | 20(55.6%) | 1(4.5%) |  |  |

## Discusstion

COVID-19 patients experience a variety of acid-base balance disorders during their course of disease. The assessment and research on the acid-base balance disorders of COVID-19 patients is still insufficient (3), which is different from our conventional understanding-the main target of damage due to COVID-19 The organ is the lung, which may cause respiratory acid-base balance disorders. A retrospective blood gas analysis study showed that the most common acid-base balance disorder in patients with COVID-19 is keratogenic alkalosis(3). A report from South Africa has been shown that metabolic alkalosis is more common in COVID-19 virus-positive patients (4). Our research also shows that metabolic alkalosis is the most common acid-base balance disorder in such patients.

The causes of metabolic alkalosis include extrarenal factors and renal factors. extrarenal factors include gastric acid loss, such as vomiting, nasogastric tube drainage, and loss of intestinal acid, such as villous adenoma, congenital celiac disease, excessive oral or parenteral intake of bicarbonate; Kidney factors such as high mineralocorticoid activity and high distal sodium delivery, persistent mineralocorticoid overdose, potassium deficiency(5).Our severe patients do not have the above-mentioned extrarenal factors, unintentional excessive intake of bicarbonate, and a small number of critically ill patients have nasogastric tube drainage, so such a high incidence of metabolic alkalosis needs to consider renal factors.

The destruction of cells entering the viral receptor, angiotensin-converting enzyme (ACE-2) II, is considered to be one of the main causes of human pathogenicity of SARS-CoV-2. ACE-2 is widely expressed in renal tubular epithelial cells, vascular components and glomerular epithelium (6). Once the SARS-CoV-2 bound ACE-2 is internalised by the cell, ACE2 is markedly downregulated (7). Theoretically, it should lead to the excessive renin-angiotensin-aldosterone system mediated by excess angiotensin II activate (8, 9).

In addition, studies have reported widespread hypokalemia in COVID-19 patients. The publication of a preprinted retrospective chinese study initially sparked interest in hypokalemia, which is a potentially common biochemical disorder in SARS-CoV-2 infection, and serum potassium was present in 108 of 175 patients <3.5 mmol/l (62%), only 10 patients had serum K> 4.0 mmol/l. Of these patients, 22% had severe hypokalemia (serum potassium <3.0 mmol/l). In total, 11% of all patients and 28% of patients with severe hypokalemia showed metabolic alkalosis (pH> 7.45), compared with 4% of patients with normal potassium. (10) However, the largest SARS-CoV-2 case series to date (including 1,099 patients) did not show any significant difference in serum potassium between mild and severe patients, in this cohort, serum potassium was mostly reported as normal (11).Our research shows that there is no significant difference in serum potassium between critically ill and critically ill patients, but both are at a low level.

Virtually all forms of metabolic alkalosis are sustained by enhanced collecting duct hydrogen ion secretion, induced by stimulation of sodium uptake through the epithelial sodium channel(12). In the renal collecting duct, mineralocorticoids drive Na+ reabsorption, K+ secretion, and H+ secretion through coordinated actions on apical and basolateral transporters(13).

Therefore, we speculate that SARS-CoV-2 uses ACE-2 as its cell receptor, leading to ACE2 degradation and ACE/ACE-2 imbalance, increasing Ang II levels, inducing the release of aldosterone and increasing mineralocorticoids, which in turn leads to blood potassium reduction and metabolic alkalosis. In addition, patients with COVID-19 often have small airway ventilatory disorders, complicated by respiratory acid. In patients with acute respiratory acidosis, PaCO2 increases by 10 mmHg, HCO3− increases by 1 mmol/l, while in chronic respiratory acidosis patients, PaCO2 increases by 1 mmol/l. In patients with acidosis, for every 10 mmHg increase in PaCO2, HCO3− increases by 4 mmol/l. In the post-hypercapnia state, respiratory acidosis improves (such as receiving mechanical ventilation), but HCO3− continues to rise, leading to metabolic alkalosis(14). In addition, in this study, the patient intake data cannot be counted in detail. Whether there is insufficient intake and aggravation of alkalosis needs further evaluation.

Metabolic alkalosis can lead to a series of serious consequences. First, elevated pH leads to respiratory depression,and alkalosis is a powerful vasoconstrictor. A large number of studies have shown that increase in pH leads to a decrease in perfusion of the heart, brain and peripheral circulation(15).

Metabolic alkalosis is the most common acid-base disorder in hospitalized patients, and it is associated with increased mortality. An earlier study by Wilson et al. in 1415 critically ill surgical patients showed that 177 (12%) developed severe metabolic alkalosis defined as arterial pH >7.54 (15). More severe metabolic alkalosis was associated with higher mortality. Mortality was 41% in patients with pH 7.55-7.56, 47% in patients with pH 7.57-7.59, 65% in patients with pH 7.60-7.64, and 80% in patients with pH 7.65-7.70. A prospective study by Anderson et al. in a group of 409 medical and surgical patients showed that mortality was 48.5% in patients with pH >7.60 (16). This study shows that among critically ill patients, the incidence of metabolic alkalosis is 100% and the mortality rate is 81.25%, and the mortality rate of severe and critically ill patients with metabolic alkalosis is as high as 95.5%.

However, the amount of data in this study is still small, and there may be therapeutic factors that interfere with the acid-base balance during the treatment of patients.If a large sample, a more detailed stratified design, and dynamic detection of patient ACE-2/renin-angiotensin-aldosterone levels can reveal more secrets.

In conclusion, in severe and critically ill patients, the proportion of metabolic alkalosis has increased significantly, and the mortality rate in patients with metabolic alkalosis has increased significantly. In COVID-19 patients, we need to pay attention to kidney damage as much as the lungs.

## Data Availability

All relevant data are within the manuscript and its Supporting Information files.

## Declarations

### Consent statement

Written consent was obtained from the patient/ guardian.

## Acknowledgments

We Thank Dr.Aiqiao Feng, Dr. Zhibin Xie, Dr Tao Li, Dr.Sai Xie, and Dr. Xiaofei Hu for contributions to the diagnosis and treatment of patients and writing suggestions.

